# TEMPORAL SPATIALIZATION AS A STRATEGY FOR URBAN MANAGEMENT IN CONTROLLING DISEASES TRANSMITTED BY MOSQUITOES

**DOI:** 10.1101/2024.02.29.24303585

**Authors:** Márlon Luiz de Almeida, Fábio Teodoro de Souza, Alzair Eduardo Pontes, Rodrigo Sant’Ana Nogueira, Raquel Ribeiro de Medeiros Baldini

## Abstract

Due to their ecophysiological ability to adapt to new environments, combined with climate change, mosquitoes that vector dengue fever and other arboviruses are increasingly expanding their areas of infestation. Dengue fever has shown growth across the planet, reaching countries and regions that were previously free of this disease. And urban management must find tools that help minimize or eradicate arbovirus vectors. This research aims to demonstrate how the temporal spatialization of diseases transmitted by mosquitoes can contribute to urban management in controlling these types of illnesses in small cities. Through a case study in a small town in the interior of Brazil, it was possible to accurately detect, through temporal spatialization, which neighborhoods were most prone to the disease from 2010 to 2020. Thus, urban management can develop public policies in these neighborhoods and improve their performance against the disease.

## Introduction

Dengue fever proves to be a global disease with disastrous consequences for health, especially in countries with tropical and subtropical climates where Aedes mosquitoes find it easy to adapt [1].

The plasticity in adapting to new environments caused the mosquitoes that transmit dengue fever (Aedes aegypti and Aedes albopictus) to spread across the globe. With climate change, there is a tendency for the vector to advance to regions that previously did not have problems with the insect [1].

The group of 17 diseases called Neglected Tropical Diseases (NTDs) – that is, with little attention to medical care, the creation of new medicines, and the formulation of diagnostic methods – are expected to have been eradicated by 2030, which includes dengue, following the Sustainable Development Goals (SDGs) [2].

Due to the urbanization processes that transform cities, there is generally an increase in population density, affecting the balance of the habitat decrease in biotic resistance concerning native species [3].

The loss of biodiversity harms Cultural Ecosystem Services (CES), which allows the entry of vector species, especially those that quickly adapt to new environments [3].

Therefore, managers need to plan and execute strategies to reduce the pandemic risk that affects the most vulnerable families as they suffer from financial scarcity. Having more incredible difficulty accessing public policies and not obtaining resources for health expenses makes them take even longer to react to combating diseases [4].

Given this scenario, this article aims to demonstrate how the temporal spatialization of diseases transmitted by mosquitoes can contribute to urban management in combating these types of illnesses in small cities, as is the case of dengue fever, for example.

It is essential to highlight that a search in the Scopus database found only 148 studies related to dengue in small cities. However, no results were found regarding the temporal spatialization of dengue in small towns.

There was a temporal spatialization study related to dengue, but it was in Guangzhou (China), a city that 2019 had around 13 million inhabitants [5]; that is, it is not tiny.

## Theoretical Reference

The traditional way of eradicating vectors using insecticides and larvicides has not had a satisfactory effect, as mosquitoes manage to develop resistance to the components of the poisons [6].

In the long term, more beneficial solutions should be planned, such as new designs and features for homes, ways to avoid water accumulation in open spaces, and improvements to the basic sanitation system, among others [6].

Dengue is the central mosquito-borne disease that most afflicts countries with tropical and subtropical climates [7]. Despite its low relative mortality, it presents high morbidity and causes financial expenditure on health by governments. Both are in an attempt to control the transmitting mosquito and are in medical care for the population infected by the virus [8].

In 2015 alone, on the American continent, there were more than two million cases, of which 1.65 million occurred in Brazil (813 points per 100 thousand inhabitants), with 811 deaths recorded [8].

The behavior of dengue fever in recent times corresponds to the disease that spreads the most throughout the world among those transmitted by mosquitoes, especially in tropical and subtropical regions. It is present in 128 countries, 36 of which are temperate regions considered free of the disease. In population terms, it is estimated that 3.97 billion people may be victims of dengue outbreaks [9]

There are approximately 390 million reports of dengue cases worldwide every year – 25% symptomatic and 1% reported with greater severity, resulting in about 9,221 deaths. Given the problems arising from the disease, the annual cost is around US$8.9 billion [9].

Due to the mosquito’s ability to adapt, dengue began to appear in some colder locations in the United States of America (USA), where it was not previously found, as in 2021, with 117 reported cases and another 513 in the territories [10], as Figure 1 suggests:

**Fig. 1.**
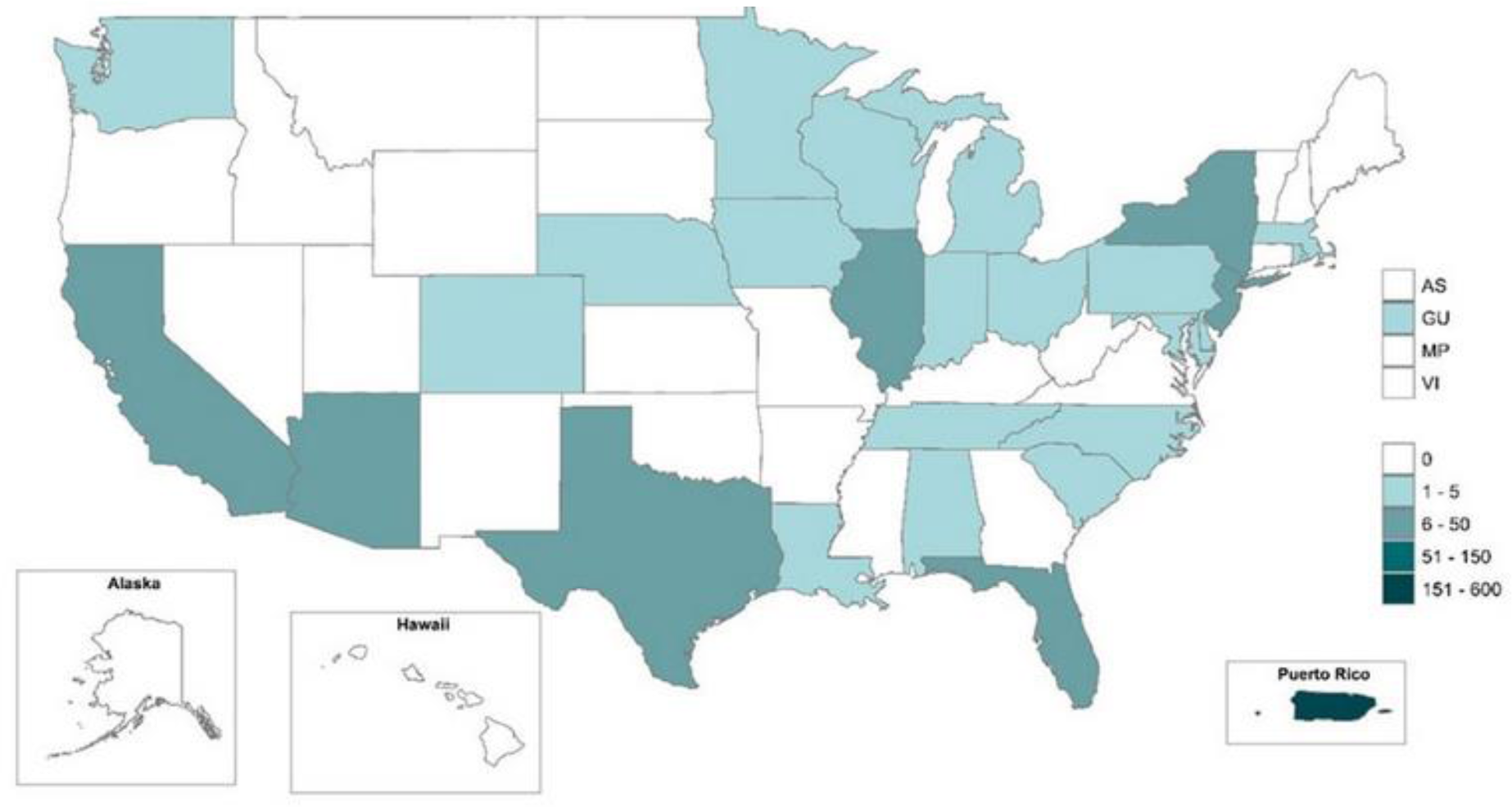
States and territories with dengue cases in the USA in 2021. Source: Centers for Disease Control and Prevention of the United States of America (2021).

The mosquito that transmits dengue fever has characteristics that make it an efficient vector. Anthropophilic in nature, it lives in the surroundings or human habitations and has versatility in its diet in just one gonotrophic cycle, increasing the chances of transmitting pathogens. Although the soaked blood complements the egg’s decomposition, it is also used as a source of energy [11].

The World Health Organization [12] explains that, until 1970, only nine countries had severe dengue epidemics, which currently occur in more than 100 nations. The increase is due to the number of cases with explosive outbreaks and the number of countries, including Europe.

In 2010, cases of the disease with local transmission were reported in France and Croatia, in addition to imported situations in three other European countries [12].

In 2012, this situation worsened with an outbreak of dengue on Madeira Island in Portugal, with more than two thousand occurrences. Consequently, cases of the disease were detected in the Portuguese nation and ten other European nations, and currently, autochthonous cases are observed in European countries [12].

The most significant dengue outbreaks occurred in 2019, when it first appeared in Afghanistan. There have been more than 3.1 million cases in the Americas, of which 25,000 were classified as severe. Other countries with high incidence were also recorded, such as Bangladesh, with 101 thousand; Malaysia, with 131 thousand; Philippines, with 420 thousand; and Vietnam, with 320 thousand [12].

In 2020, several countries reported an increase in the number of cases, namely Bangladesh, Brazil, Cook Islands, Ecuador, India, Indonesia, Maldives, Mauritania, Mayotte, belonging to France, Nepal, Singapore, Sri Lanka, Sudan, Thailand, Timor -East and Yemen. In 2021, dengue plagued Brazil, India, Vietnam, the Philippines, the Cook Islands, Colombia, Fiji, Kenya, Paraguay, Peru, and the Reunion Islands [12].

Improvements in the quality of life of populations in developing countries cannot accompany the expectation of doubling the urban growth rate by 2050. It must be stated that the increase in cases of arboviruses in the world was practically due to urbanization and environmental deterioration, poverty, and social inequality [13].

Pandemics (and their consequences) should lead cities to review concepts about service provision, as assessed [14], and rethink how they plan their space.

In other words, when urban management begins to understand and know the community’s changing needs, it provides conditions to develop planning integrated with public services based on the community and implement solutions that connect local governments, community associations, and the population [14].

However, there is a need to create urban management strategies to mitigate the effects of a pandemic on low-income families (vulnerable populations) [4].

These authors consider that economic vulnerability, difficulty accessing public policies, and the minimum capacity to spend on health can interfere with the municipality’s ability to react with policies to combat disease outbreaks.

Economic and social vulnerability is a complicating factor for urban management, as it spatially concentrates several families in the exact location more susceptible to the effects of a pandemic [15].

The dengue is transmitted by Aedes mosquitoes, which reproduce quickly in environments with poor water and sanitation conditions. It is necessary to involve specialized state mobilization that leads the population to an adaptive capacity, understanding the need for environmental and social contingencies that improve human well-being; as such, families understand the ecological changes [15].

Challenges and problems in cities – restriction of environmental resources, congestion, air pollution, and emission of greenhouse gases, among others – require, from governments and people, forms of urban management that use technologies and innovations [16].

In the case of dengue transmission, which follows characteristic seasonal patterns within the framework of a year and is cyclical, it presents disease outbreaks in specific years; creating a database with information about the disease and its causes is essential for its control. [17].

It should be noted that NTDs are diseases linked to poverty and cannot be classified as diseases of developing nations. Their effects are felt by economically vulnerable communities in all parts of the planet, even if people with less financial means live in rich countries [18].

## Materials and Methods

The methodological proposal of this research was to carry out a case study of the incidence of dengue in a small city in the interior of Brazil through the temporal spatialization of confirmed cases of the disease in the city by neighborhood.

The case study systematically investigates an event or a set of related events that propose to describe and explain the phenomenon of interest. Although the case study is mainly used for prospective studies, it is also widely used for retrospective studies [19].

### 3.1 Characterization of the municipality researched in the case study

Goiatuba is located in the state of Goiás, in the Central-West region of Brazil. It has 35,649 inhabitants and a population density of 14.38 inhabitants/km2 and is situated at an altitude of 720 m.

The average income of formal workers is 2.5 minimum wages (in Brazil) per month (around USD 675.00), with 28.9% of its population having some occupation. The percentage of people with a per capita income of up to half the minimum wage is 30.9% [20].

In turn, the Municipal Human Development Index (IDHM) is 0.725, while the afforestation of public roads is at 87.6%, and urbanization at 29.6% [20] – the Gini Index corresponds to 0.5176 [21].

The water network is 211,480 m long and serves 13,495 connections (99.9%), and the sanitary sewage network is 103,000 m, serving 6,540 links (54% of the total). Sewage is not treated. In 2020, the Municipal Development Index (IDM) was 5.28 (general) and 2.88 (infrastructure) [22].

### 3.2 Database

Regarding data on the disease mentioned in this work, the Historical Series of Dengue Cases Notified by Epidemiological Week was used, provided by the Health Surveillance Superintendence (SUVISA) of the Goiás State Department of Health.

Also, from SUVISA, the Notification/Investigation Bulletin was obtained, such as Frequency per Epidemiological Week with Notification and Classification (Dengue) and Frequency per Epidemiological Week Notification and Evolution (Deaths), belonging to the Aedes Zero Integrated Monitoring System.

The Epidemiological Surveillance Centers of municipalities, states, and the Federal District register notifications and investigations based on the epidemiological calendar of the epidemiological weeks of each year, which the Federal Government defines on the Sinan website. With this, each week’s start and end dates are determined without coinciding with the weeks of the regular annual calendar [23].

The Brazilian territory comprises municipalities with varied characteristics in area, population, socio-environmental and economic specifications, and altitude, among others. It is worth noting that public data on diseases, outbreaks, epidemics, and pandemics from the Federal Government are generally recorded based on what occurs in each location. They can be considered at the level of street, neighborhood, or area of the city and are georeferenced [24].

The Geographic Information System (GIS) offers possibilities for using cognition to understand spatial configurations and perceptions, especially from computer mapping and spatial analysis [25].

Integrating GIS as support and prediction for decision-making in dynamic geographic processes, such as emergencies and disasters, is increasingly common. Still, it allows future scenarios to be shown based on specific situations [25].

GIS can also be used to construct spatial statistics to reduce the effects of disease outbreaks based on scientific information and to understand the “propagation dynamics,” considered the most relevant property of an epidemic or outbreak [26].

For this study, the application of GIS was based on georeferenced data in the State Geoinformation System of the State Department of Health of the State of Goiás, spatial information from Google Maps (2022) and ArcGIS Datum SIRGAS 2000 – UTM Coordinate System 22S.

Maps of the researched city were created and divided into neighborhoods. In each one, the number of dengue cases in each year (from 2010 to 2020) was superimposed, with the identification of each quantity through colored circles that varied in size proportional to the number of cases.

Finally, a map was created that encompasses all years of the research (2010 to 2020) with overlapping periods to check whether dengue cases were more recurrent in any neighborhood at the time and try to identify a spatio-temporal pattern of recurrence.

## Results and discussion

GIS to locate reported cases of dengue in Goiatuba (GO) allows the identification of the respective neighborhood where the disease occurred and in each year from 2010 to 2020. Furthermore, it favors verifying the location where dengue is most recurrent and a history of recurrence [24–25].

Creating maps of Goiatuba and identifying dengue cases by neighborhood makes it possible to understand whether there is a dynamic spread of the disease [26].

In Figures 02 to 13, the map of Goiatuba (GO) is presented, referring to the period from 2010 to 2020 and the dengue cases by neighborhood. They are identified by colored circles and proportional to the number of reported disease cases.

**Fig. 02.**
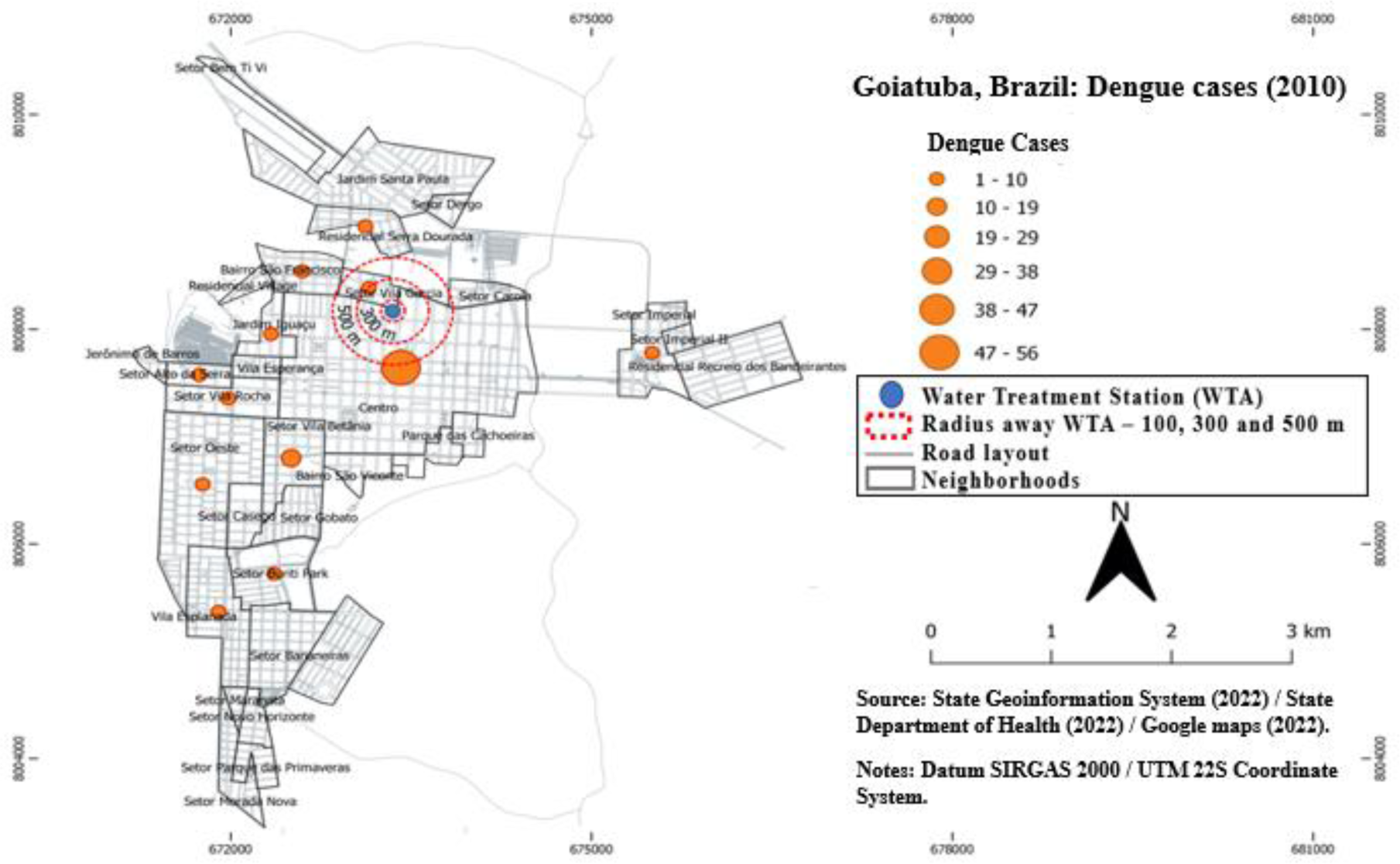
Dengue cases in Gudauta, Brazil – 2010 Source: Prepared by the authors (2023).

**Fig. 03.**
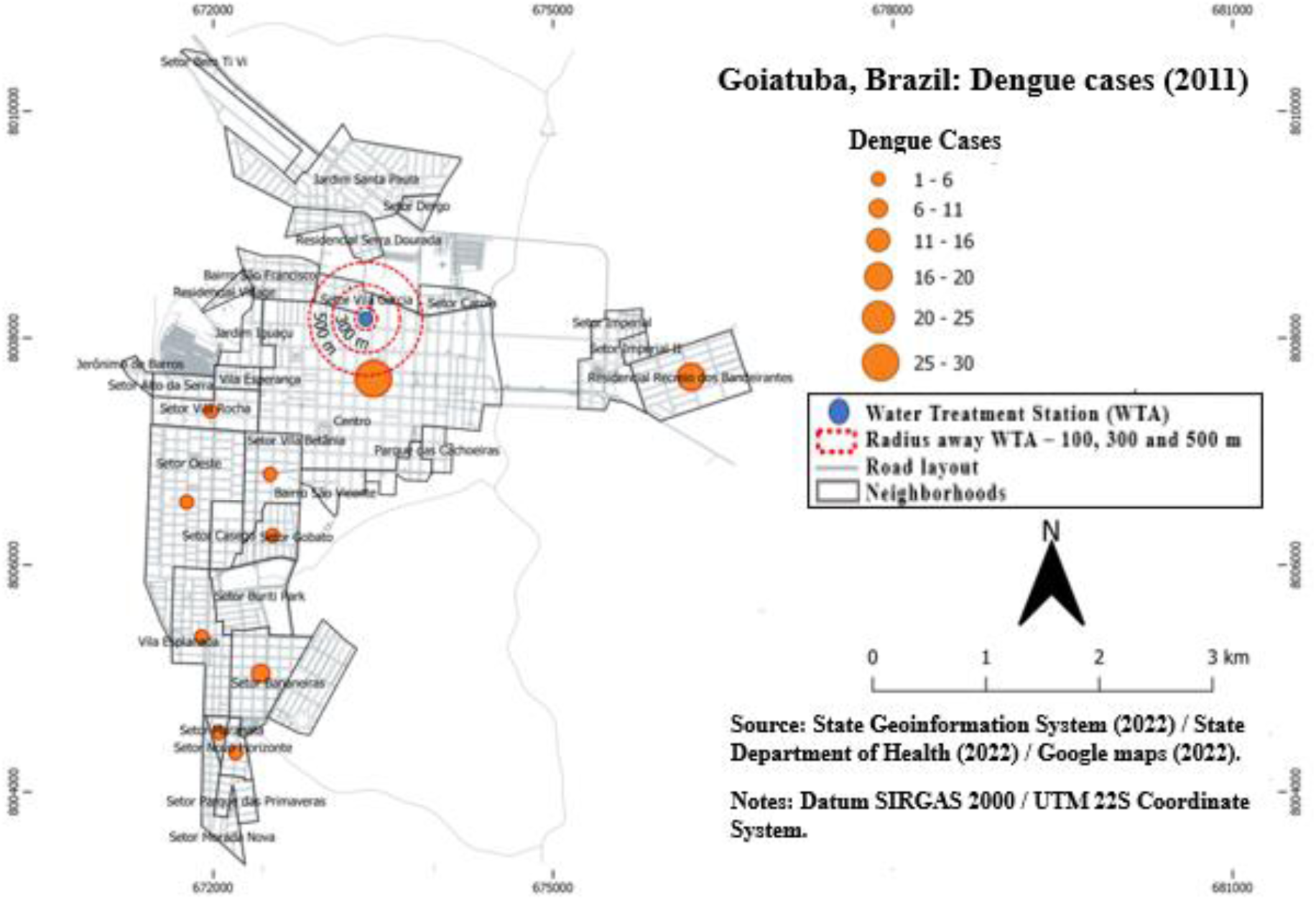
Dengue cases in Goiatuba, Brazil – 2011 Source: Prepared by the authors (2023).

**Fig. 04.**
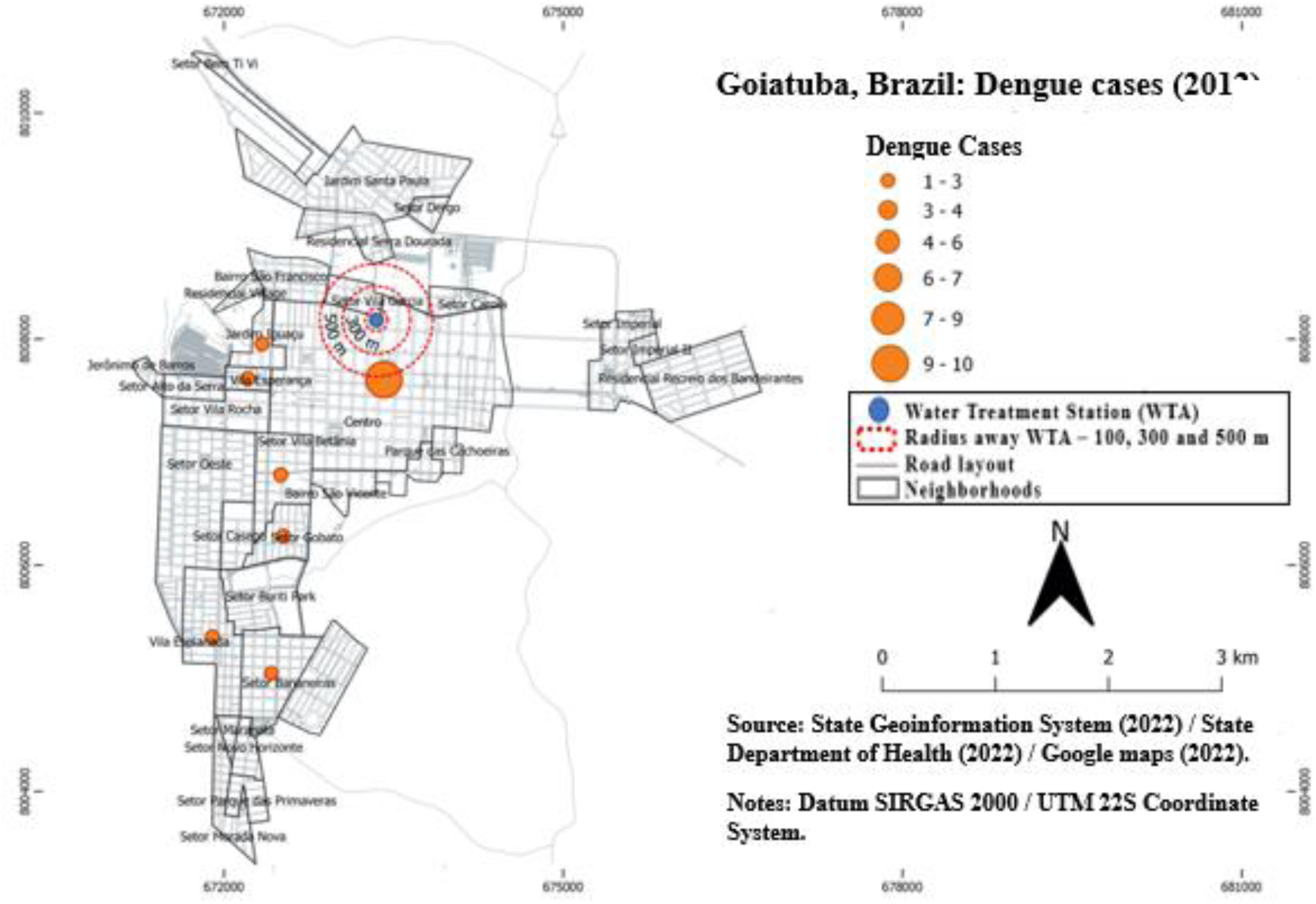
Dengue cases in Goiatuba, Brazil – 2012 Source: Prepared by the authors (2023).

**Fig. 05.**
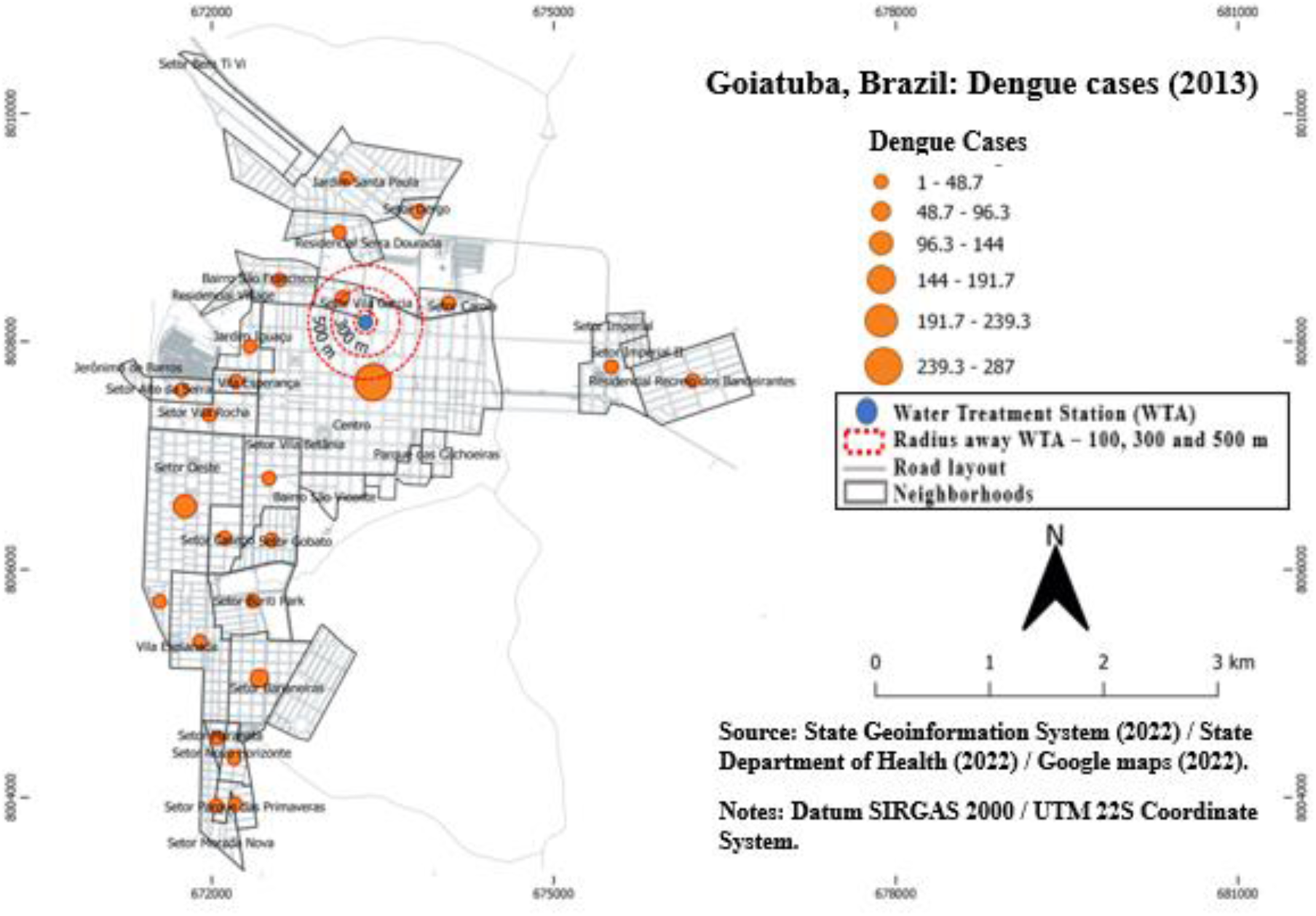
Dengue cases in Goiatuba, Brazil – 2013 Source: Prepared by the authors (2023).

**Fig. 06.**
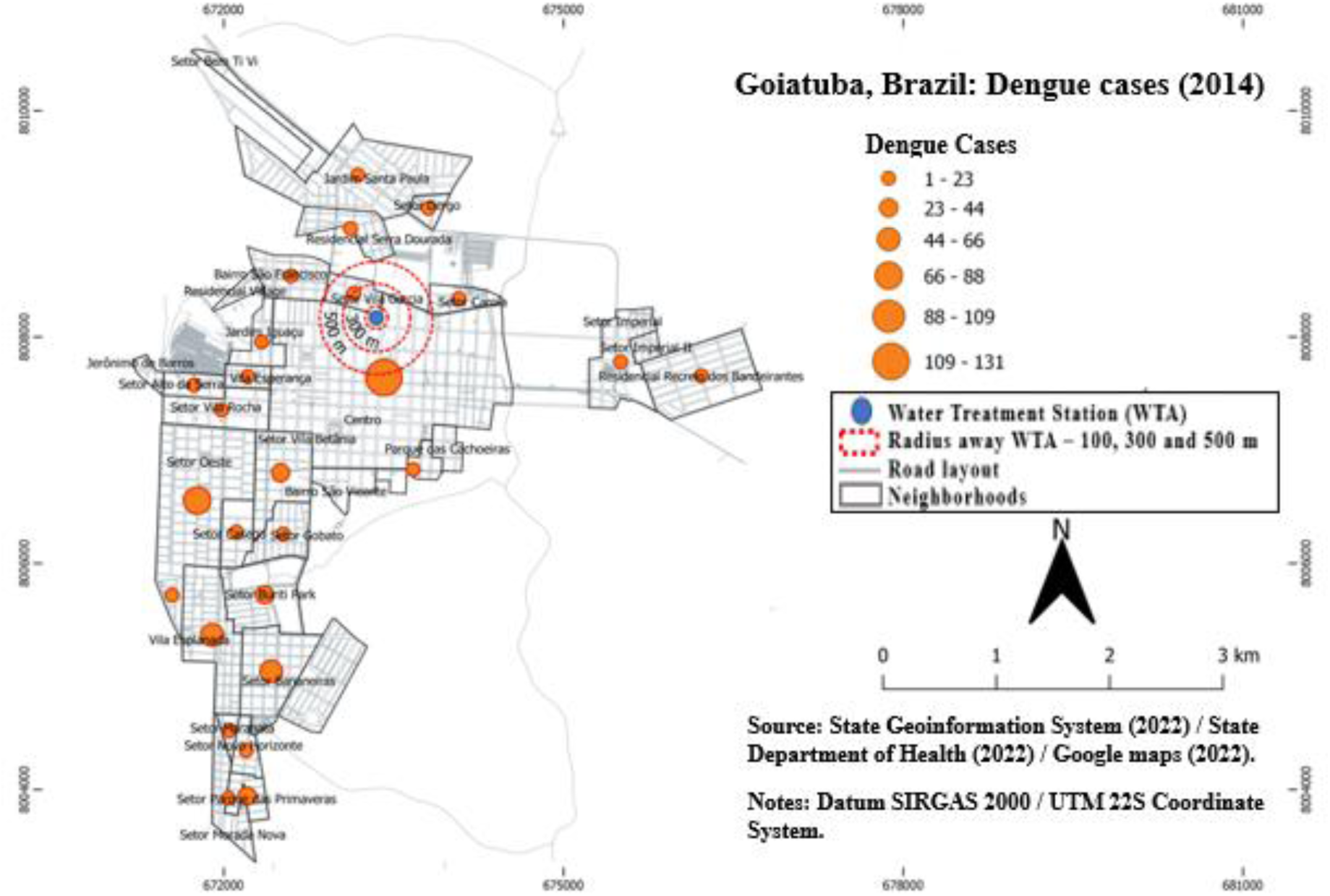
Dengue cases in Goiatuba, Brazil – 2014 Source: Prepared by the authors (2023).

**Fig. 07.**
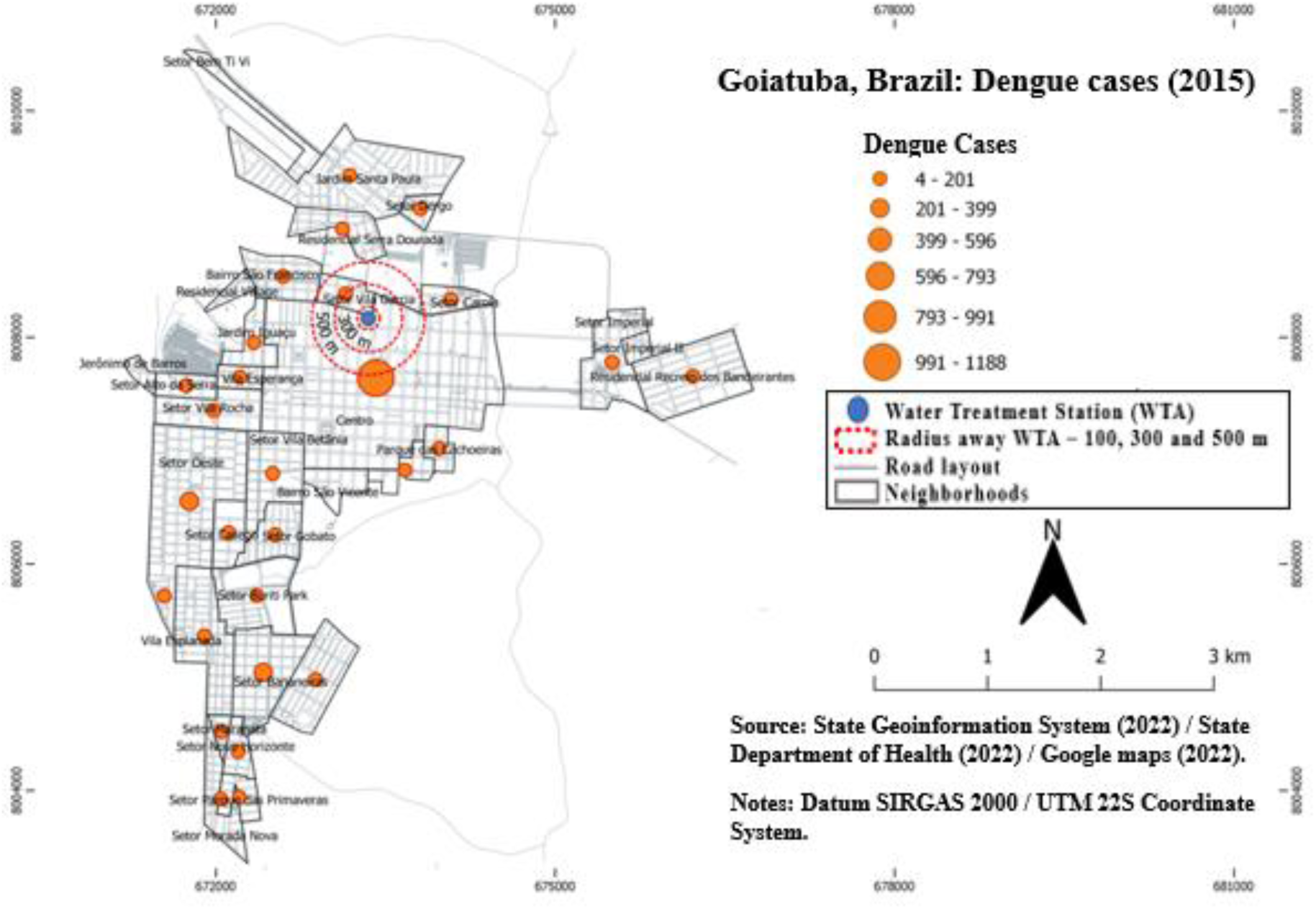
Dengue cases in Goiatuba, Brazil – 2015 Source: Prepared by the authors (2023).

**Fig. 08.**
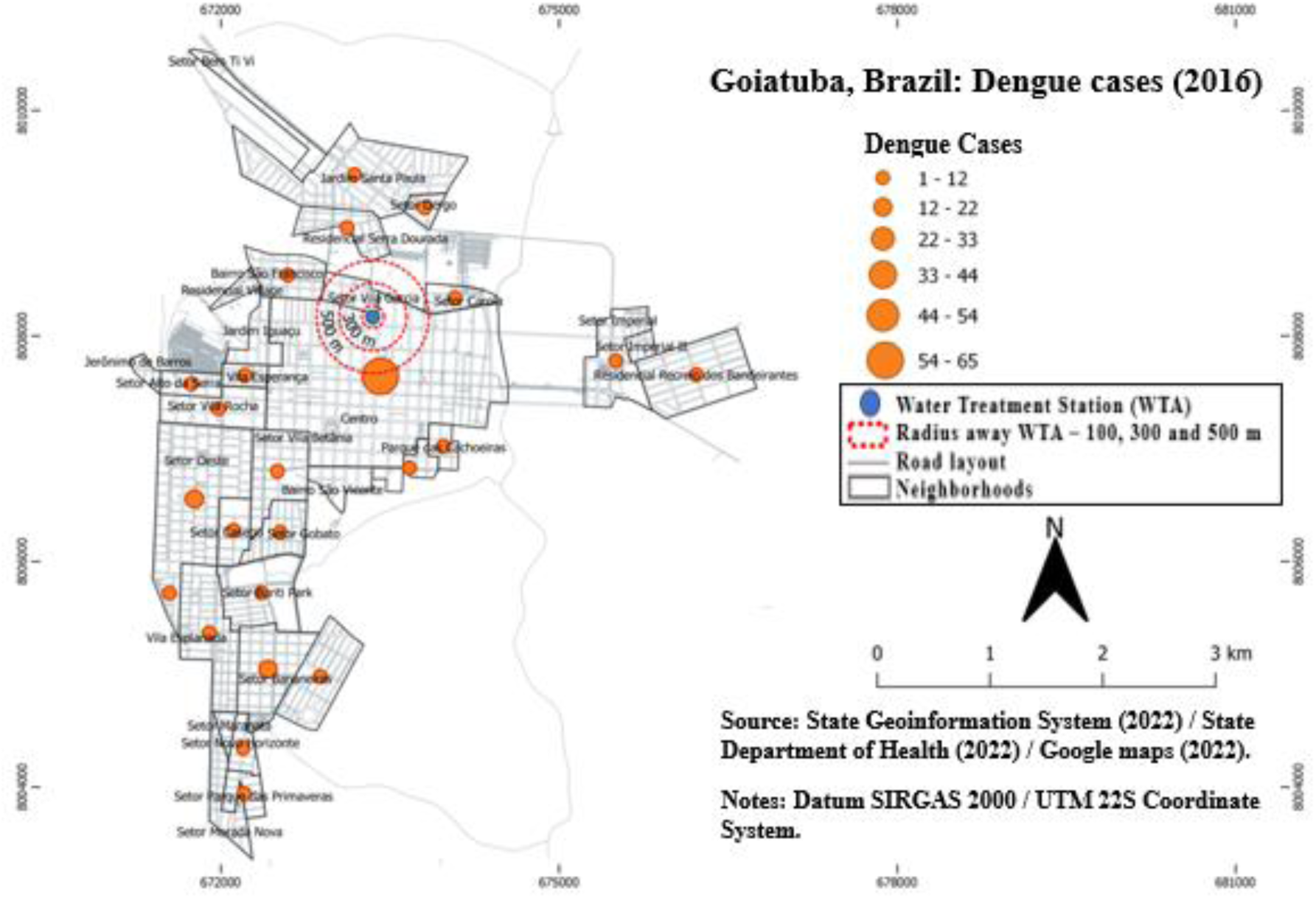
Dengue cases in Goiatuba, Brazil – 2016 Source: Prepared by the authors (2023).

**Fig. 09.**
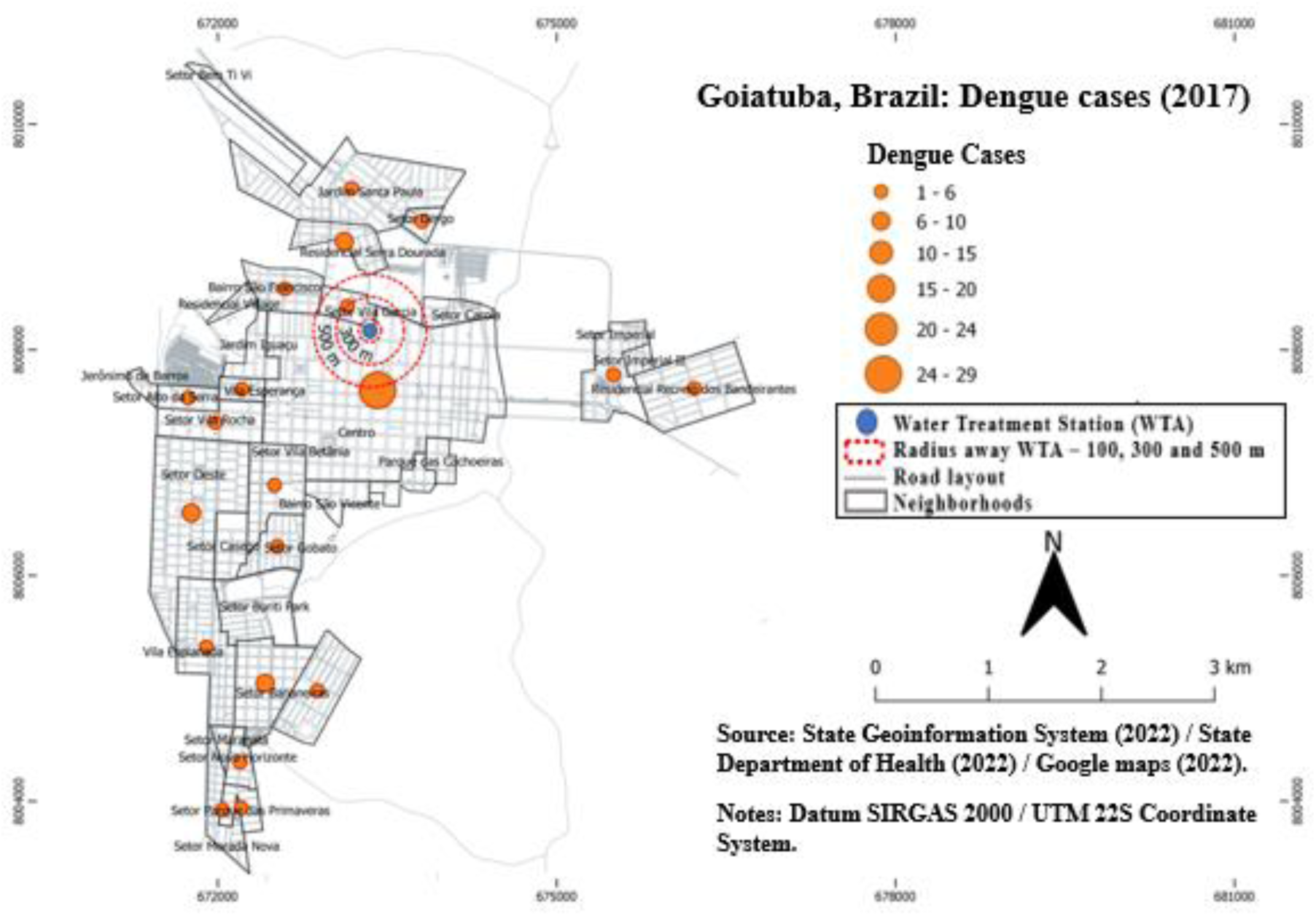
Dengue cases in Goiatuba, Brazil – 2017 Source: Prepared by the authors (2023).

**Fig. 10.**
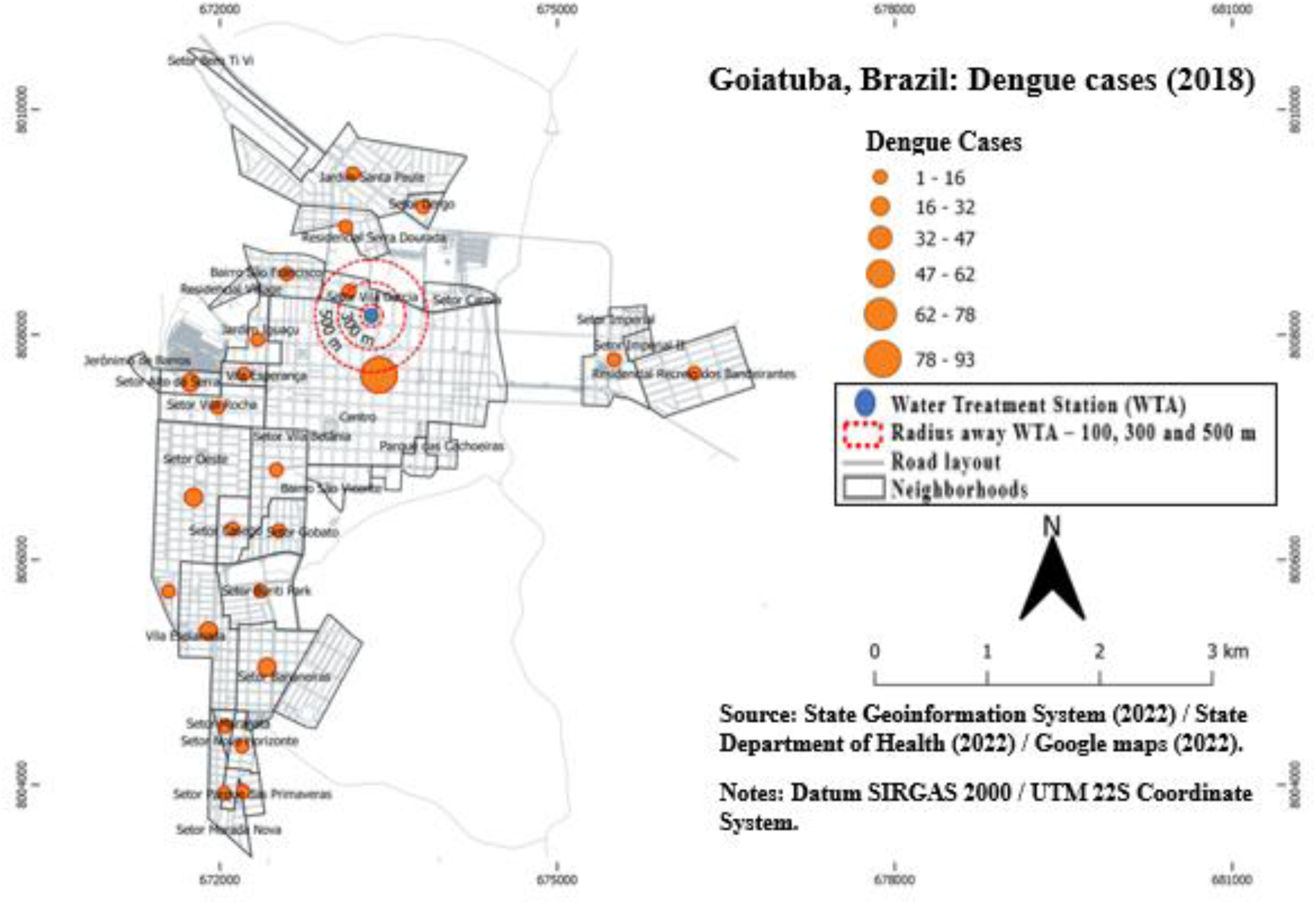
Dengue cases in Goiatuba, Brazil – 2018 Source: Prepared by the authors (2023).

**Fig. 11.**
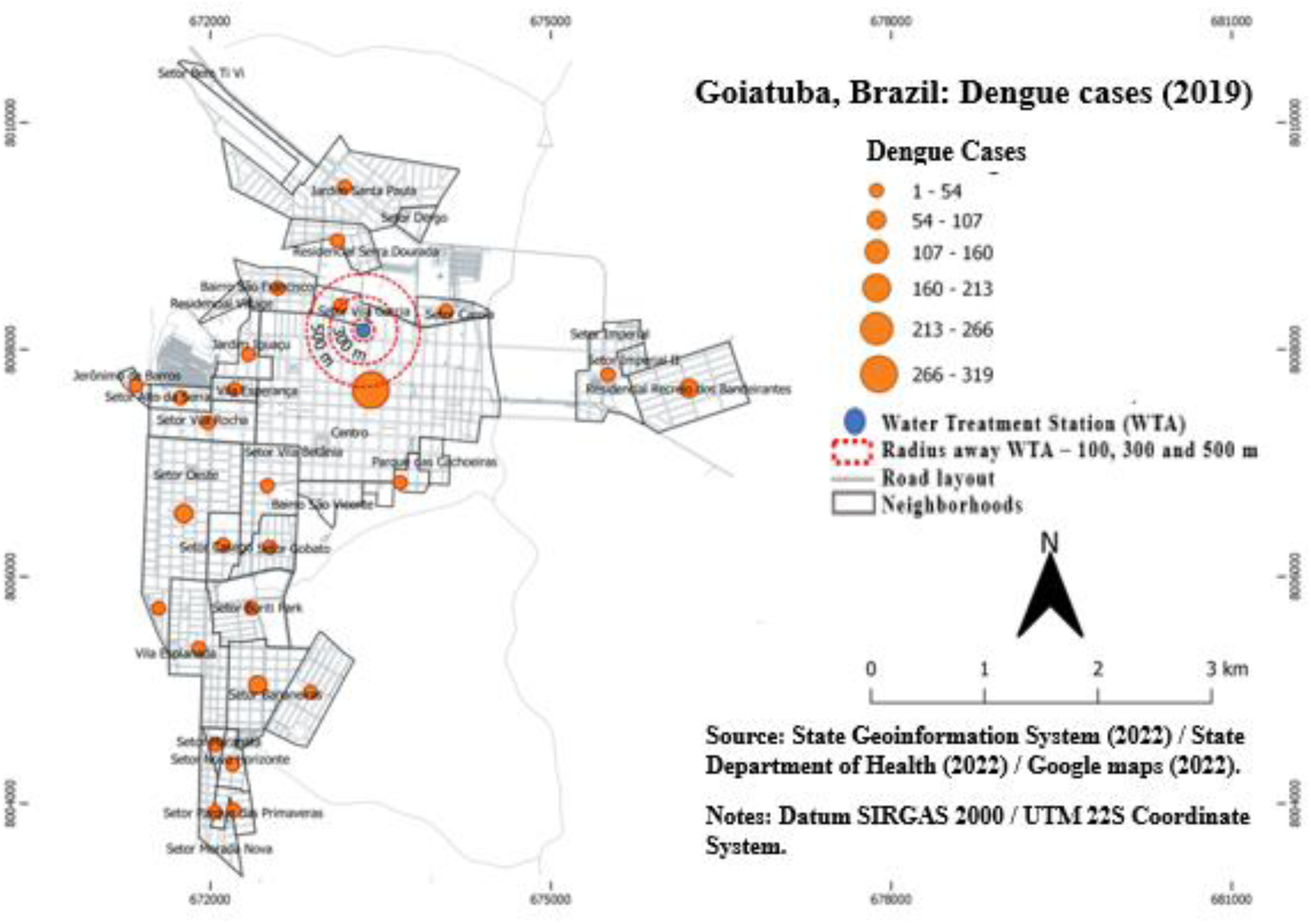
Dengue cases in Goiatuba, Brazil – 2019 Source: Prepared by the authors (2023).

**Fig. 12.**
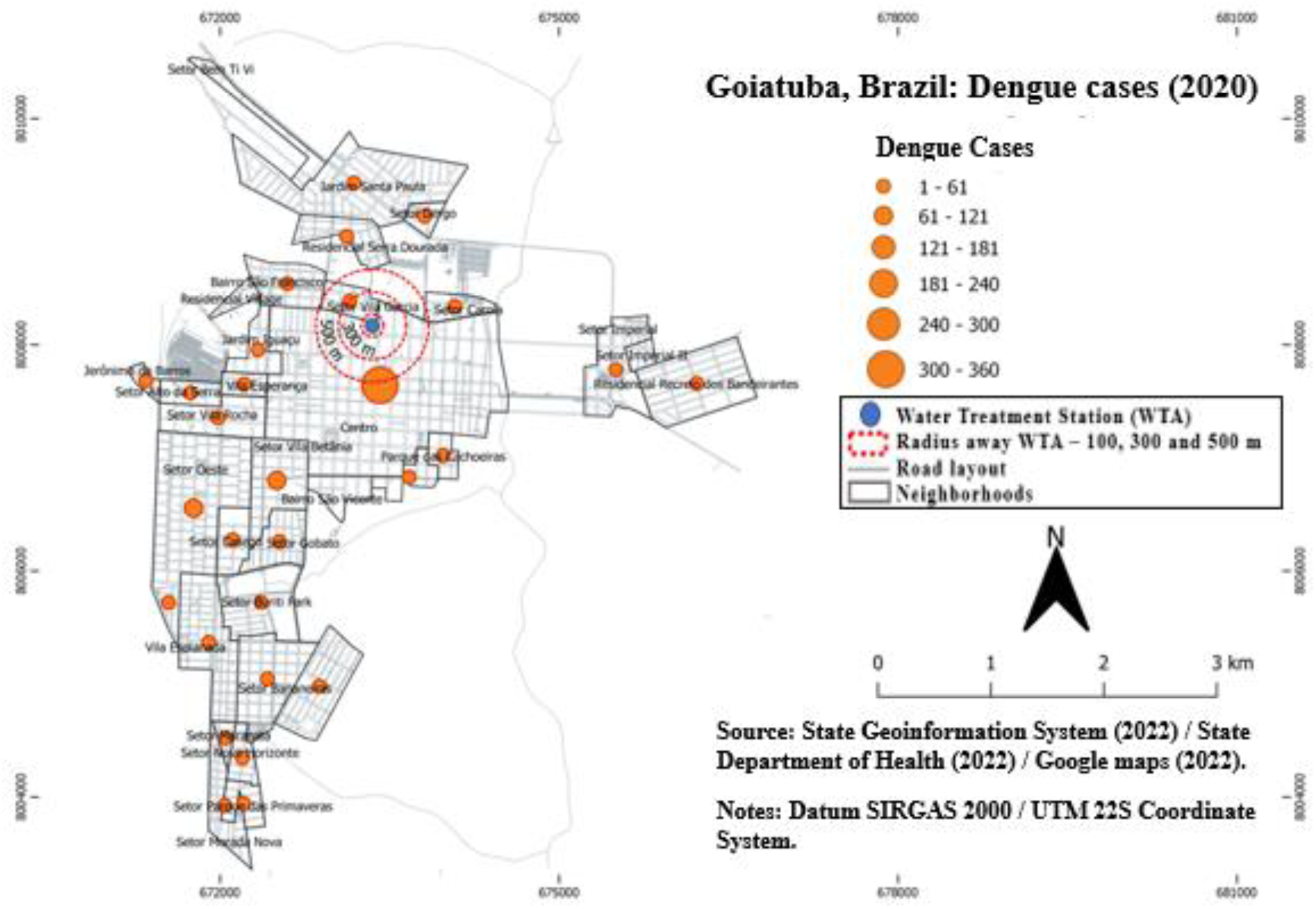
Dengue cases in Goiatuba, Brazil – 2020 Source: Prepared by the authors (2023).

**Fig. 13.**
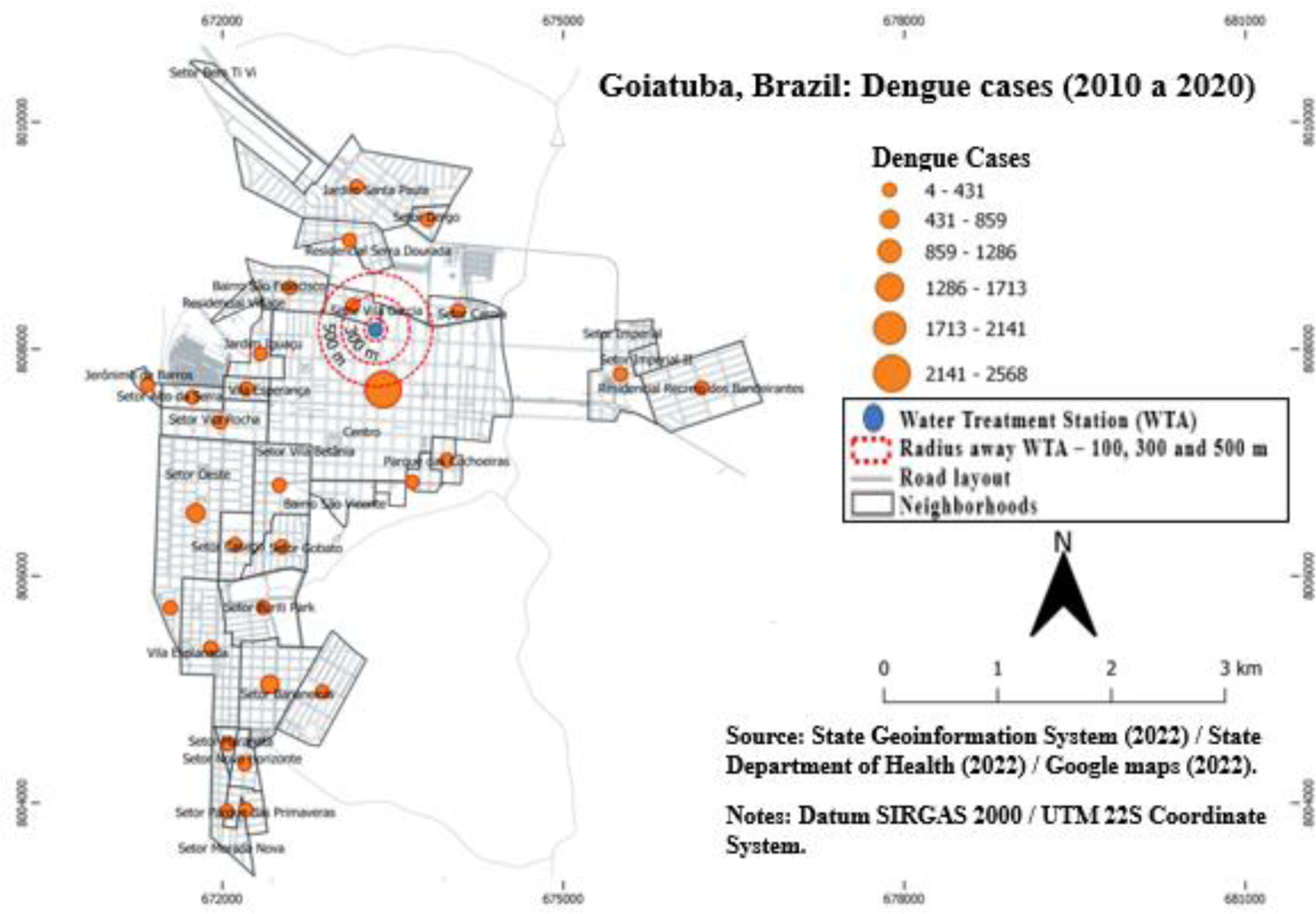
Dengue cases in Goiatuba, Brazil – 2010 a 2020 Source: Prepared by the authors (2023).

In the work of mapping reported cases of dengue by neighborhood in Goiatuba, Brazil, analyzed using the GIS, in addition to allowing a clear view of where the disease manifested itself in a more intense or mild form, the software enables creating graphs with the trend of dengue fever disease, according to the period under study.

Figure 14 illustrates the trend in dengue behavior in Goiatuba, Brazil. The trend line for dengue cases was drawn by the software that created the GIS maps based on the issues reported and identified in the city.

**Fig. 14.**
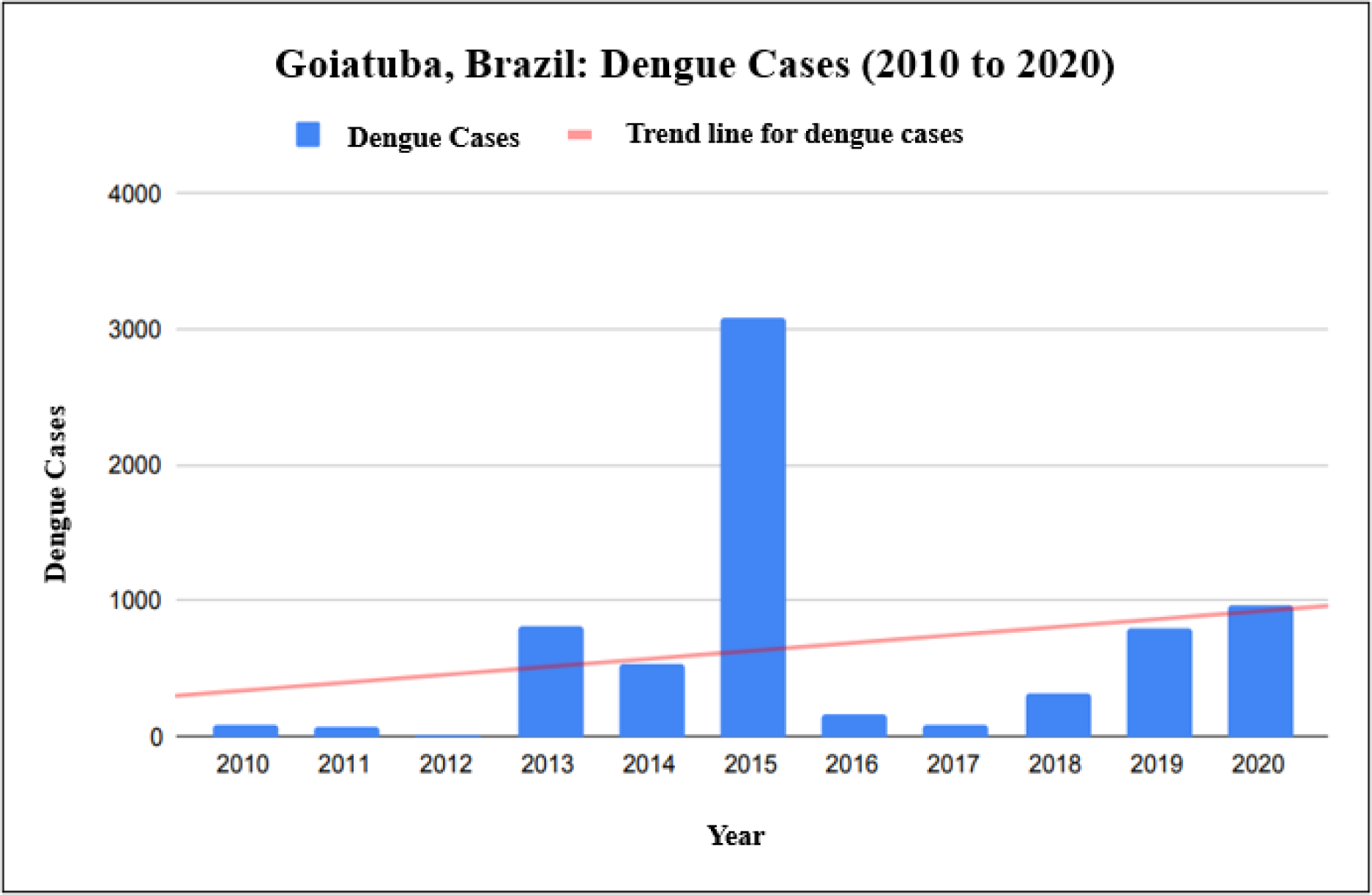
Trend for dengue cases – Goiatuba, Brazil – 2010 to 2020 Source: Prepared by the authors (2023).

In the graph and its respective trend line for dengue cases, it can be seen that Goiatuba, Brazil, shows a prospect of growth for the disease.

In this sense, it becomes crucial to understand whether the neighborhoods with the highest number of reported cases are peripheral or central and whether there is economic-social vulnerability since this aspect, according to the approach of [15], makes management work more complex urban, as there are families susceptible to the disease.

Identifying neighborhoods where dengue is recurrent can collaborate with urban management actions to understand the strategies and public policies that can obtain better results, especially regarding offering the safety of essential domestic utilities [4], such as water, energy, and community access to health infrastructure.

The solution involves the search for solutions that can connect local managers with their respective populations to rethink the plans for occupying their spaces [14].

Even though dengue appears in practically every neighborhood in the city, urban management needs to be aware that some places tend to be more effective in the emergence of cases of the disease.

From these locations, there is passive dispersion of the mosquito vector in each neighborhood, sometimes even far from each other, provided by the help of people from the city. This aspect is favored by the peculiar interactions between viruses, vectors, hosts, and the environment [27].

Using the comparison maps of reported dengue cases by neighborhood, it was possible to check where the highest number of disease incidences were reported from 2010 to 2020 and establish a ranking from first to fifth place.

In this way, it is possible to demonstrate whether some locations are more recurrent than others, that is, whether any location is more susceptible to the disease. Therefore, urban management can investigate the behavior and dynamics of dengue in the city.

When establishing the ranking of neighborhoods that reported the highest number of dengue notifications, Table 1 shows the classification of such locations in the city researched:

**Table 01.** Classification of areas in Goiatuba-Brazil, about the number of dengue cases – 2010 to 2020.

| Year | 1st place | 2nd place | 3rd place | 4th place | 5th place |
| --- | --- | --- | --- | --- | --- |
| 2010 | Center | Betânia Village | San Francisco | Esplanade I | West Sector |
| 2011 | Center | Recreio dos Bandeirantes | Bananeiras Sector | West Sector | Betânia Village |
| 2012 | Center | Esplanade II | Gobato Sector | Iguaçu Garden | Bananeiras Sector |
| 2013 | Center | West Sector | Bananeiras Sector | Betânia Village | Recreio dos Bandeirantes |
| 2014 | Center | West Sector | Bananeiras Sector | Parque das Primaveras | Buriti Park |
| 2015 | Center | West Sector | Bananeiras Sector | Recreio dos Bandeirantes | Betânia Village |
| 2016 | Center | West Sector | Bananeiras Sector | Recreio dos Bandeirantes | Betânia Village and Rocha Village |
| 2017 | Center | West Sector | Bananeiras Sector | Serra Dourada | Recreio dos Bandeirantes |
| 2018 | Center | Bananeiras Sector | West Sector | Esplanade II | Recreio dos Bandeirantes |
| **2019** | **Center** | **Bananeiras Sector** | **West Sector** | **Recreio dos Bandeirantes** | **New Horizon** |
| **2020** | **Center** | **West Sector** | **Betânia Village** | **Recreio dos Bandeirantes** | **Esplanade I** |

Source: Prepared by the authors (2023).

In Table 01, the neighborhood of Goiatuba, Brazil, most frequently affected by dengue infestation was the Center – in fact, this sector was the highlight from 2010 to 2020, as it came in first place every year.

Then, of the 11 possible years, the West Sector was in second place on six occasions; and, in third place, Bananeiras Sector can be chosen, which, twice, was in second place and, on six occasions, was in third place. In addition, it is necessary to pay attention to Recreio dos Bandeirantes.

It is known that the dengue vector mosquito has the capacity for adaptation reported by [1] and [10] and for its impressive reproductive ease, including the transmission of the virus to its descendants by the infected female [12].

It is postulated that urban management must be attentive to the dynamics of the disease in the city, especially in the neighborhoods where it is most recurrent, as detected in this research in Goiatuba, Brazil.

Even though meteorological elements influence dengue cases, the fact that cases of the disease are not uniform for all neighborhoods in the city leads to uncertainties that cannot be explained solely concerning meteorological aspects.

They must be investigated by urban management to find the element that most favors the emergence and increase of cases in specific locations.

From the point of view of this investigation, which sought to point out strategic ways to face, combat, and minimize the effects of dengue, the identification of neighborhoods with the highest incidence of the disease does not mean that urban management can only use corrective actions, such as the application insecticides or other control actions after the onset of the disease.

Such subsequent mechanisms of action do not bring the expected effects and may even promote the emergence of more resistant mosquitoes [6].

Therefore, it is necessary to continue monitoring the dengue vector and verify the vulnerabilities that can lead to the emergence of outbreaks when mapping the distribution of disease cases in Goiatuba, Brazil.

Therefore, the anthropic factors that enhance the multiplication of *Aedes* mosquitoes in these locations must be observed.

## Conclusion

The task of eradicating the 17 NTDs by 2030 as a prerequisite for the UN SDGs, which includes dengue, is initially a significant challenge, especially for small cities in developing countries. To the detriment of climate change and the ecophysiological capacity of mosquitoes that transmit the disease, this disease has increasingly spread across the planet and in regions where it was previously not even thought it could exist.

Faced with the imminent dengue pandemics worldwide, mainly in countries with a tropical climate (but not exclusively), public authorities must seek mechanisms to confront, control and eradicate the dengue vector, as without a vector, there is no disease. Such tools can be translated into public health policies to ensure the population’s quality of life and well-being.

However, for public policies to be efficient, practical, and effective, their execution must be based on some instruments or tools that contribute to urban management in the adequate planning of such policies.

This way, the result can reach the expected goal, and the cost-benefit ratio becomes economical for the city administration.

It is worth noting that temporal spatialization was managed in this research to identify the neighborhoods of the cities with the highest incidence of confirmed dengue cases without stating whether these neighborhoods were also the most infested with mosquitoes that transmit the disease.

In this sense, future research should seek to clarify the mosquito vector infestation rate in neighborhoods with the highest number of cases of the disease, even to understand whether there is a relationship between the number of patients and the infestation rate.

## Data Availability

Data on dengue cases by street and neighborhood in Goiatuba, state of Goiás, Brazil, can be obtained from the Superintendence of Health Surveillance (SUVISA) of the Department of Health of the State of Goiás at the email address “.”

